# Variability in SARS-Cov-2 IgG Antibody Affinity To Omicron and Delta Variants in Convalescent and Community mRNA Vaccinated Individuals

**DOI:** 10.1101/2022.03.01.22271665

**Authors:** Michael K. Tu, Samantha H. Chiang, David T.W. Wong, Charles M Strom

## Abstract

The emergence of Omicron and Delta variants of SARS-CoV-2 has begun a number of discussions regarding breakthrough infection, waning immunity, need and timing for vaccine boosters and whether existing mRNA vaccines for the wildtype strain are adequate. Our work leverages a biosensor-based technique to evaluate the binding efficacy of SARS-CoV-2 S1 specific salivary antibodies to the Omicron and Delta variants using a cohort of mRNA vaccinated (n=109) and convalescent (n=19) subjects. We discovered a wide range of binding efficacies to the variant strains, with a mean reduction of 60.5%, 26.7%, and 14.7% in measurable signal to the Omicron strain and 13.4%, 2.4%, and −6.4% percent mean reduction to the Delta Variant for convalescent, Pfizer, and Moderna vaccinated groups respectively. This assay may be an important tool in determining susceptibility to infection or need for booster immunization as the pandemic evolves.

**Key Points:**

- AMPERIAL assay developed to quantify salivary SARS-CoV-2 S1 IgG antibodies to Omicron and Delta variants
- There was a reduction in affinity to both Delta and Omicron Variants
- The reduction in affinity was more pronounced to Omicron than for Delta Variants
- There was a significant difference between IgG affinities in Individuals vaccinated with Pfizer versus Moderna Vaccines

## INTRODUCTION

As the COVID-19 epidemic continues into its third year, two major variants (first the Delta and subsequently the Omicron) have become the predominant strains in infected individuals. The two FDA approved mRNA vaccines are based on wild-type sequences of the Spike protein. With the emergence of the Omicron variant, some fully vaccinated individuals (including those who had a third booster injection) were still susceptible to COVID infection. The susceptibility to infection could be the result of waning immunity, a reduction in affinity of vaccine induced antibodies to the Omicron S1 protein, or a combination of the two. Questions regarding susceptibility to infection and long term immunity are being actively investigated (1) as well as speculation regarding the relationship between natural immunity and vaccine-induced antibody production (2).

Our laboratory has previously developed a non-invasive saliva-based quantitative biosensor assay for Anti-S1 IgG antibody levels to the wild-type SARS-CoV-2 using a proprietary platform called AMPERIAL™. We demonstrated that this assay is highly specific to COVID-19 infection (3) and can be used to serially monitor vaccinated patients for waning antibody levels (4). As the COVID-19 pandemic continues with a number of variants being monitored to determine their potential harm and differences in transmission rate, a major point of interest is whether existing mRNA vaccines based on the wild-type S1 sequence would remain efficacious with successive new variants.

The Amperial™ platform allows rapid development of quantitative assays for antibodies to SARS-CoV-2 variants. As soon as variant S1 antigen becomes available commercially, it can be immobilized in the conducting gel and used to quantify IgG levels in saliva. In the present study, we developed quantitative Amperial™ assays to both Delta and Omicron S1 antigen. We then simultaneously measured IgG levels to the wild-type and Variant S1 antigens in a cohort of over 100 Pfizer or Moderna vaccinated individuals and 19 convalescent patients who contracted COVID-19 prior to the emergence of either the Delta or Omicron variants. We found a significant reduction in antibody affinity for the Delta variant and Omicron variants in all 3 cohorts with affinity to Omicron more reduced than to Delta. We also found that Pfizer vaccinated individuals have a lower affinity to the Omicron variant than Moderna vaccinated individuals. Our work also indicates that antibodies derived from natural immunity have reduced reactivity to the Omicron variant relative to the wildtype or Delta variant.

## MATERIALS AND METHOD

### SARS-CoV-2 Salivary Quantitative Antibody Assay

As previously described, the Amperial® platform uses an integrated electrode and electrochemical reader system (EZLife Bio Inc, Los Angeles, CA) to read the oxidation-reduction byproduct of the antibody capture process. The detailed description of the Amperial® COVID-19 Antibody assay and the assay performance and validation have been described previously (3). We reported the utility of this assay in monitoring the kinetics of antibody response (4) using immobilized S1 subunit to the wild-type virus immobilized in a conducting gel on the surface of the gold electrode.

We selected the S1 antigen as the capture antibody because both the Pfizer and Moderna vaccines use mRNA coding for the S1 antigen. In our work we evaluated the difference between wildtype, omicron, and delta variants through the immobilization of the identical concentrations of Delta and Omicron S1 antigen and electrochemically measuring the captured SARS-CoV-2 S1 specific antibodies present in saliva. For the delta variant, we used SARS-CoV-2 variant S1 Antigen B.1.6.617.2 (40591-V08H23 SinoBiological, Wayne, PA), a recombinant antigen that included T194, G142D, E156G, 157-158 deletion, and the L452R, T478K, D614G, 681R mutations. For the Omicron variant, we used SARS-CoV-2 variant S1 Antigen B.1.1.529 (40591-V08H41), a recombinant antigen with the stated mutations of “A67V, HV69-70 deletion, T95I, G142D, VYY143-145 deletion, N211 deletion, L212I, ins214EPE, G339D, S371L, S373P, S375F, K417N, N440K, G446S, S477N, T478K, E484A, Q493R, G496S, Q498R, N501Y, Y505H, T547K, D614G, H655Y, N679K, P681H”(5).

Equal concentrations of the wildtype, omicron, or delta S1 antigen were immobilized in separate gold electrodes (0.3 µg/well) on a 96-well electrochemical plate via an electropolymerized polypyrrole. Following coating of the SARS-CoV-2 antigen on the surface with the conducting polymer, salivary samples were diluted 1:10 in Casein/PBS and incubated on the plate for 10 minutes. Following a PBS-T wash, biotinylated anti-human IgG Fc is incubated on the plate as a secondary antibody, washed off with PBS-T, and then a horseradish peroxidase enzyme chain is incubated and washed off. The final measurement is performed by adding a H2O_2_/TMB solution and performing chronoamperometric measurement at −200 mV at 60 seconds to measure the final electrochemical current generated by the completed assay complex. For the evaluation of SAR-CoV-2 efficacy, each experimental run consisted of wells where each sample was pipetted on a surface coated with wildtype antigen, variant antigen (Omicron or Delta antigen), and a polymer only well which was used to normalize for background effects.

### Human Subjects

Research protocol and consents were approved by the Western Internal Review Board (Study #1302611). Vaccinated individuals under the age of 18 years, receiving immunosuppressive drugs, had prior COVID-19 infections, or received cancer chemotherapy, were excluded from the study.

Volunteers who had previously received a Pfizer (BioNTech) or Moderna for SARS-CoV-2 were consented. Subjects were issued a questionnaire collecting information about vaccination dates and vaccine type, along with questions to eliminate subjects who met exclusionary criteria. Saliva samples were collected using the Orasure® Oral Fluid Collection Device (Orasure Technologies, Bethlehem, PA). Individuals with documented COVID-19 infection were also consented and samples obtained.

We analyzed saliva samples from a cohort of fully vaccinated individuals with Pfizer or Moderna mRNA vaccines in addition to a set of unvaccinated convalescent patients. The detailed demographics of the patients appears in table 1. The existing set of specimens is generally weighted towards the population that is 60+ years old. All samples were received prior to the emergence of the Omicron variant. We performed simultaneous analyses for SARS-CoV-2 specific antibodies using identical amounts of either the wild-type or Omicron S1 protein immobilized in a conducting gel in the Amperial™ assay^2,3^. The percentage difference in measured current between wild-type and Omicron antigen was calculated for each individual.

**Table 1.**
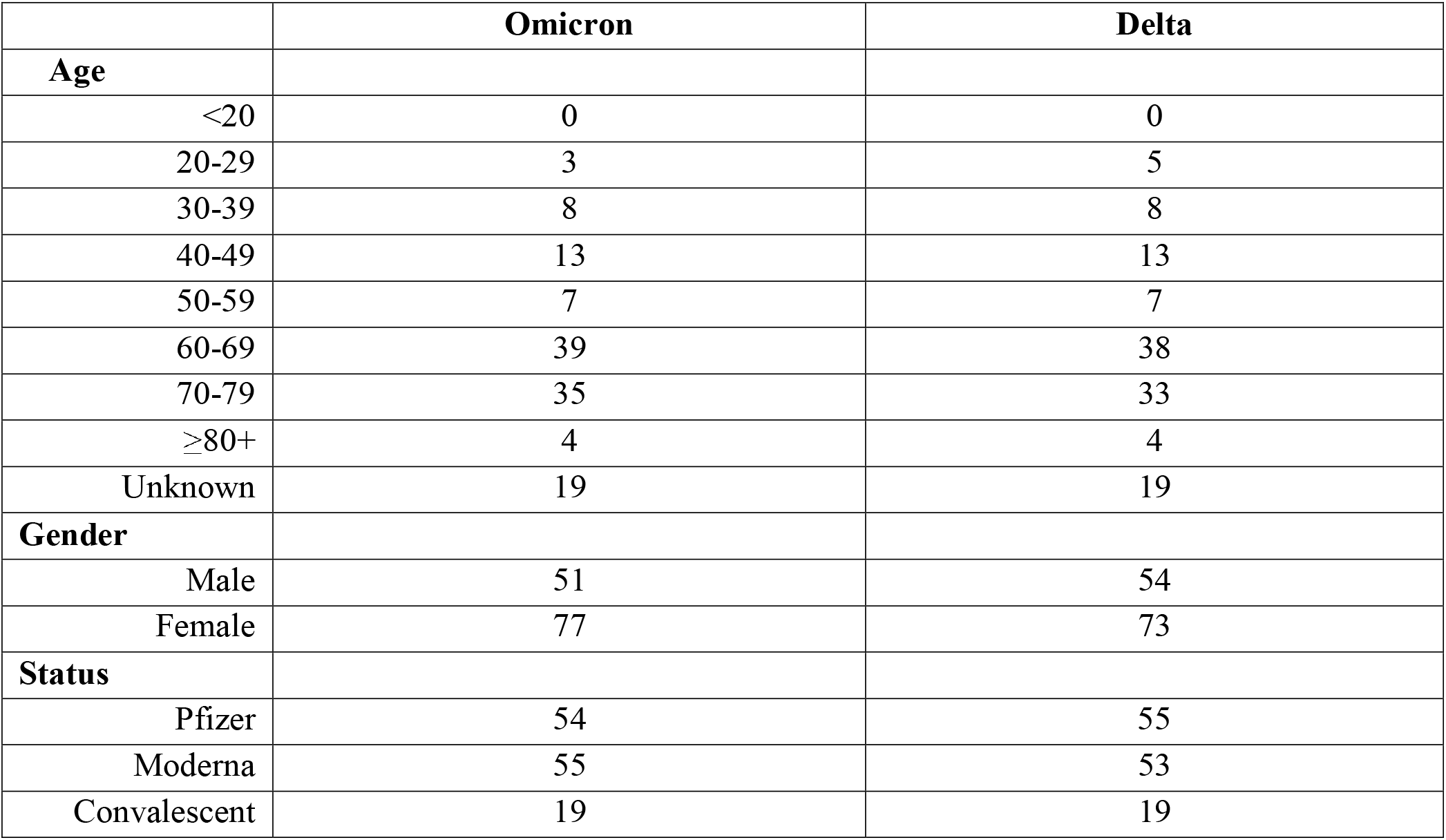
Summary demographics of vaccinated cohort.

|  | Omicron | Delta |
| --- | --- | --- |
| **Age** |  |  |
| <20 | 0 | 0 |
| 20-29 | 3 | 5 |
| 30-39 | 8 | 8 |
| 40-49 | 13 | 13 |
| 50-59 | 7 | 7 |
| 60-69 | 39 | 38 |
| 70-79 | 35 | 33 |
| ≥80+ | 4 | 4 |
| Unknown | 19 | 19 |
| **Gender** |  |  |
| Male | 51 | 54 |
| Female | 77 | 73 |
| **Status** |  |  |
| Pfizer | 54 | 55 |
| Moderna | 55 | 53 |
| Convalescent | 19 | 19 |

## RESULTS

### Affinity to Delta and Omicron Variants

Available samples were tested using the AMPERIAL™ assay, with each plate containing a wild-type in comparison with a variant antigen (omicron or delta) for each sample. Following completion of each experiment, comparisons were calculated between the measured current of the wildtype on each plate and the variant antigen on each plate. For the omicron antigen, 55 Moderna, 54 Pfizer, and 19 convalescent samples were tested. For the delta antigen, 53 Moderna, 55 Pfizer, and 19 convalescent samples were tested. We then calculated a percent reduction using the antibody level to the wildtype minus the antibody level to the variant divided by the antibody level to the wild-type antigen. This calculation yields the percentage of reduction in affinity to the variant (Figure 1).

**Figure 1.**
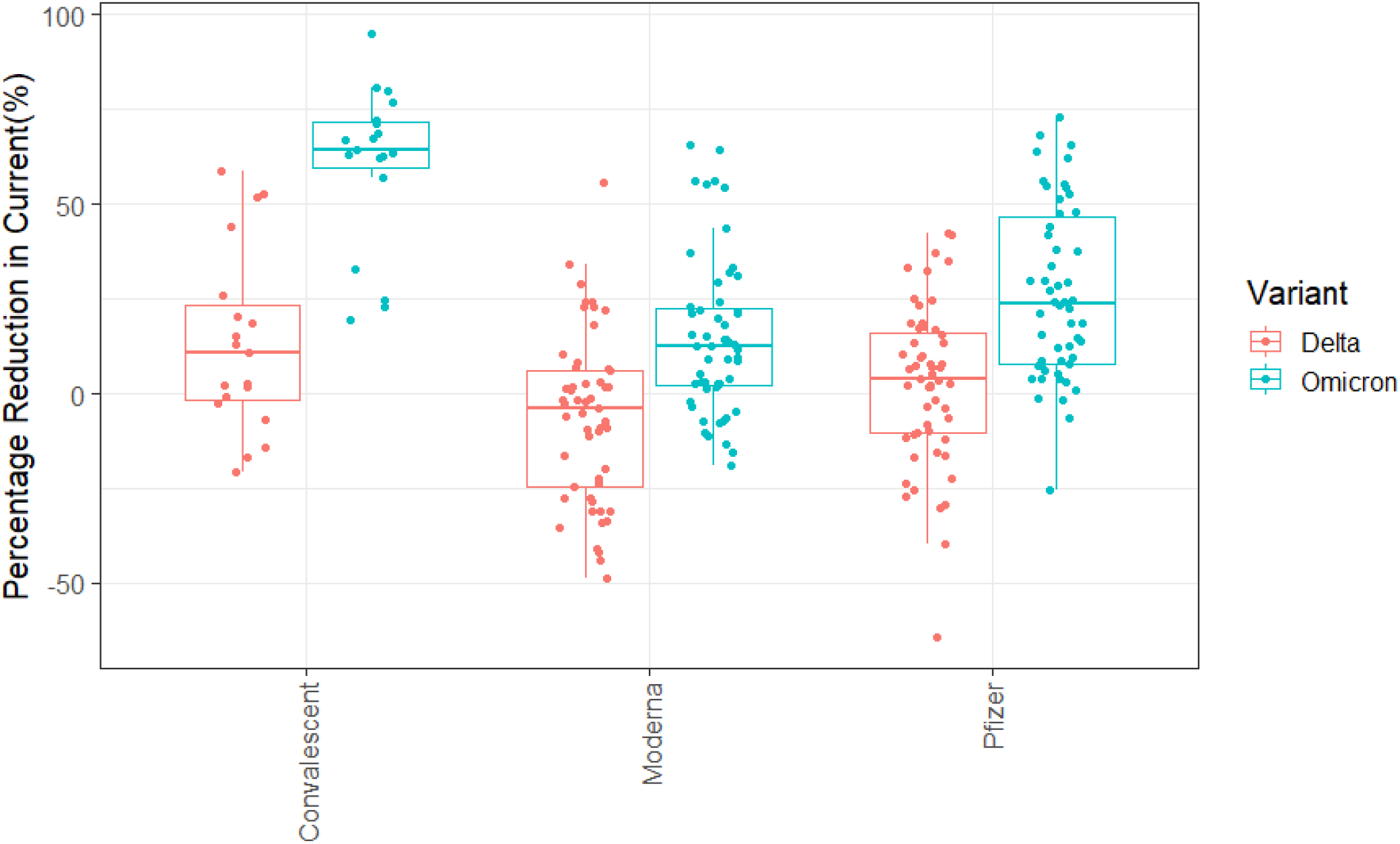
Percentage reduction relative to wildtype for samples run for Delta and Omicron variants of SARS-CoV-2 for different categories.

**Figure 2.**
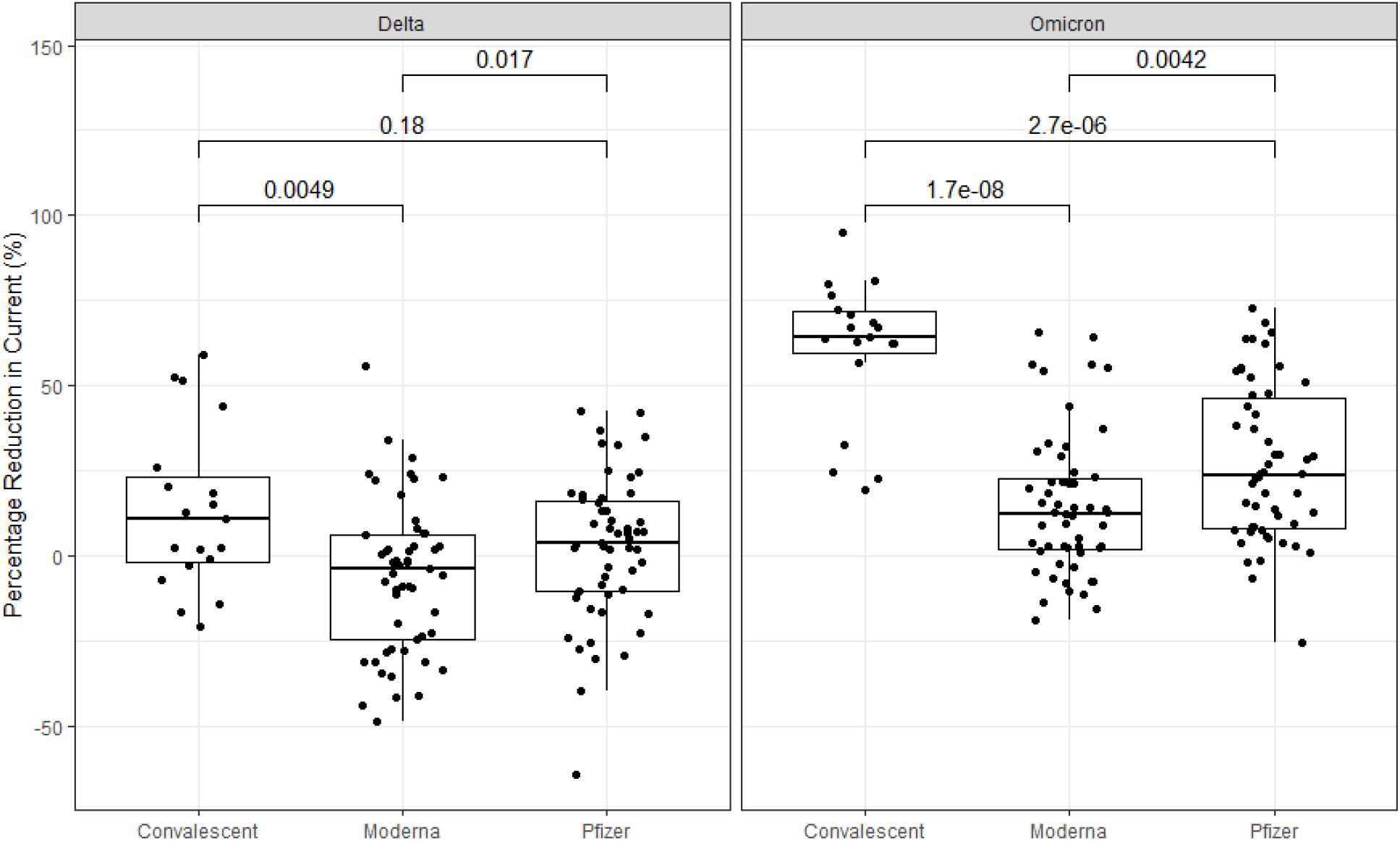
Within variant comparison of antibody level reduction.

**Figure 3.**
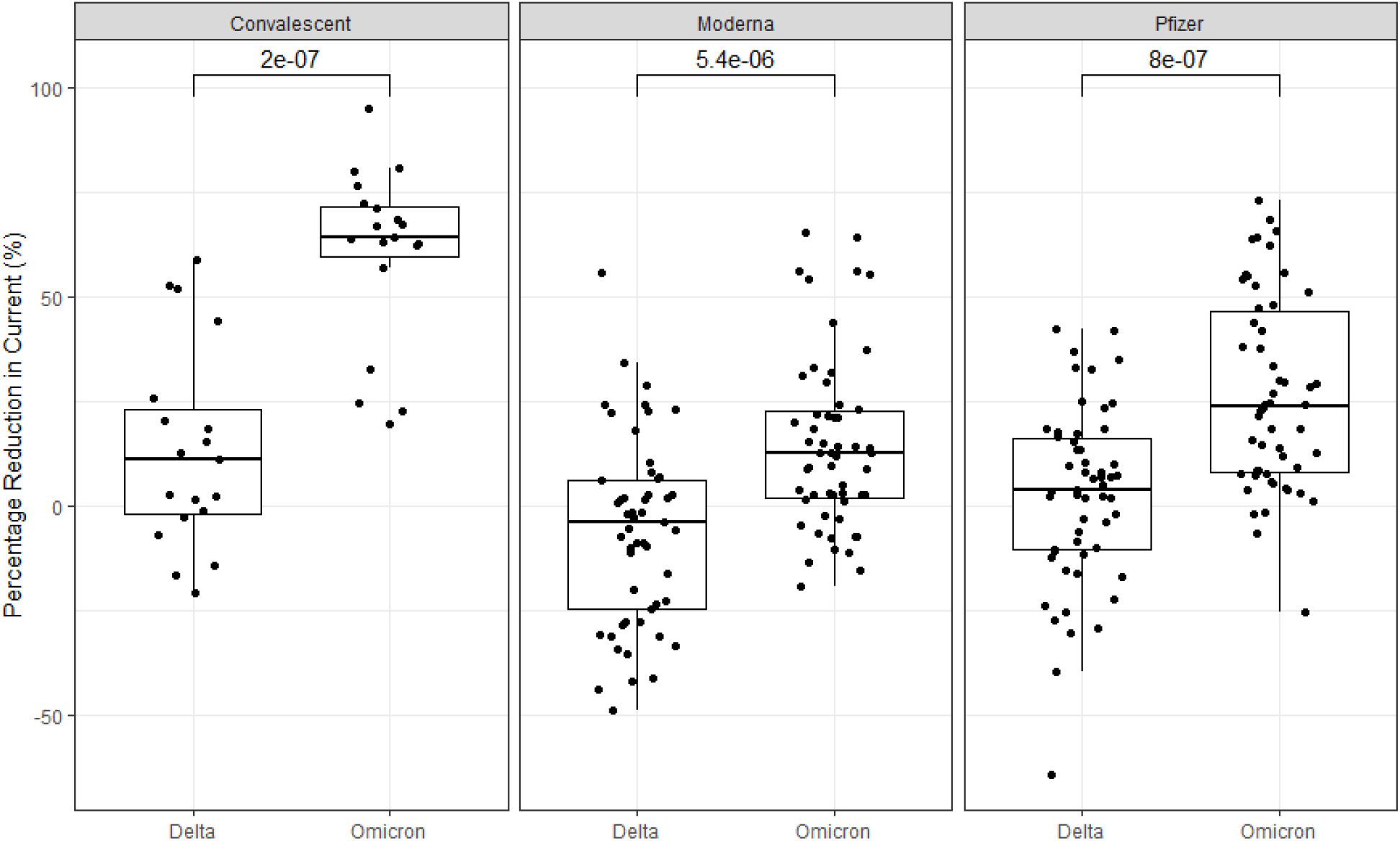
Within antibody group comparison of antibody level reductions

Analysis of the different groups showed that for the omicron variant there were reductions of 60.5%, 26.7%, and 14.7% in mean signal compared to wildtype for the convalescent, Pfizer, and Moderna samples respectively. For the delta variant, a 13.4%, 2.4%, and −6.4% percent mean reduction to the delta variant was observed for convalescent, Pfizer, and Moderna samples respectively. There was a wide variation in affinities among individuals. Many individuals had little reduction or even increased affinity to the variants, whereas others had significant reduction of 50% or more. The reductions were greatest in the convalescent patients, followed by the omicron and then the delta variants.

We performed paired sample paired t-tests for each individual sample run with both the wild-type and the variant assays. Within the Omicron variant group, statistical significance was found between the Pfizer and Moderna vaccines (p=0.0042), the Pfizer and Convalescent group (p=2.7e-06), and the Moderna and Convalescent group (p=1.7e-08). Within the Delta variant group, statistical significance was found between the Pfizer and Moderna vaccines (p=0.017) and the Moderna and Convalescent group (p=0.0049) while there was no significance found between the Pfizer and Convalescent group (p=0.18).

For comparisons within the same category of antibodies: within the convalescent group, statistical significance was found between the omicron and delta variants (p=2e-07). Within the Moderna group, significance was found between omicron and delta variants (p=5.4e-06). Within the Pfizer group, significance was found between the omicron and delta variants (p=8e-07).

To summarize, statistical significance was found in all cases of comparisons within variant groups and within antibody groups excluding the case between Pfizer and Convalescent antibodies for the Delta variant.

Table 2 is summary of the percentage of individuals in each group that experience a >50% reduction in antibody affinity to the variant. Of note is that in the convalescent group, 3 individuals (15%) had a >50% reduction in affinity to the Delta variant and 13 of 19 individuals (79%) had a >50% reduction in antibody affinity to the Omicron variant. In the vaccinated cohorts, 22% of Pfizer vaccinated individuals experienced a greater than 50% reduction in antibody affinity to the omicron variant and 11% of Moderna vaccinated individuals had similar decreases.

**Table 2:**
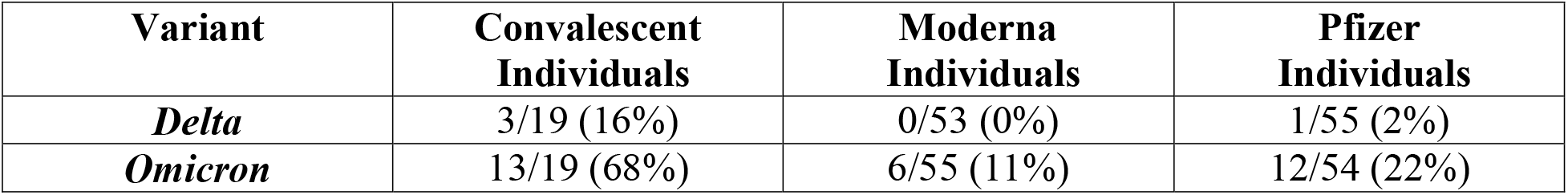
Percentage of individuals experiencing a >50% drop in affinity to SARS-CoV-2 Variants

| **Variant** | **Convalescent**<br>**Individuals** | **Moderna**<br>**Individuals** | **Pfizer**<br>**Individuals** |
| --- | --- | --- | --- |
| ***Delta*** | 3/19 (16%) | 0/53 (0%) | 1/55 (2%) |
| ***Omicron*** | 13/19 (68%) | 6/55 (11%) | 12/54 (22%) |

## DISCUSSION

The data presented demonstrate the omicron variant provides a greater risk and susceptibility compared to wildtype or delta variants of SARS-CoV-2. As the experimental results show, there is a significant decrease in binding affinity (14.7%-26.7%) for the Omicron variant relative to the wildtype, and there are significant differences between all groups of delta and omicron variants, suggesting a greater risk of susceptibility to the Omicron variant than the Delta variant. The reduction in antibody affinity is highly variable among individuals. More than 50% show little or no reduction in affinity to the Omicron variant whereas in some groups over 20% experience a >50% reduction in affinity. This may explain the somewhat haphazard susceptibility to COVID-19 among vaccinated individuals. Our data also demonstrate that individuals vaccinated with the Moderna vaccine have lower incidences of affinity reduction to the Omicron variant. There is no guarantee that the next variant will show the same differences. Perhaps a “mix and match” scenario where individuals receive boosters of a brand different from their original vaccine may be a possible strategy for inducing a broad range of protection.

In this study specifically we only evaluated the affinity of IgG antibodies to the S1 subunit of the spike protein of SARS-CoV-2, and consequently only serves as an evaluation of a subset of the antibodies related to immunogenicity. Nevertheless, inasmuch as the S1 subunit is the component coded for by the existing mRNA vaccines, the results herein presented still offer a large degree of insight into the reduced efficacy of antibodies in light of the various mutations present in the variant strains.

There was a wide variety of antibody affinity to the omicron variant among individuals, with some having equivalent binding while others had close to a 70% reduction in binding efficacy. Although currently there is no proven method to predict susceptibility to COVID-19 infection using antibody measurement, it is likely that a combination of decreasing antibody levels with decreased affinity of antibodies to the omicron variant are contributing to the current spike in COVID-19 infections among vaccinated individuals.

Our data suggests that as more variants emerge, a modification to the existing mRNA vaccines may be required to prevent infection and transmission. By serially measuring both antibody levels and antibody affinity we may be able to determine appropriate timing of booster vaccinations.

While managing a global pandemic typically involves a large variety of assessments beyond functional antibodies, this study adds to the mounting evidence that salivary antibody assessment is a non-invasive, scalable, and viable method of evaluating for the dynamics of immunity during a pandemic. Since saliva measurement can be done without the involvement of health care professionals, this platform can allow broad based testing of entire populations in order to inform decision making.

## Data Availability

All data produced in the present study are available upon reasonable request to the authors

## Acknowledgments

The authors wish to thank our volunteers who participated in the study.

